# Wastewater detection of emerging arbovirus infections: Case study of Dengue in the United States

**DOI:** 10.1101/2023.10.27.23297694

**Authors:** Marlene K. Wolfe, Abigail Harvey Paulos, Alessandro Zulli, Dorothea Duong, Bridgette Shelden, Bradley J. White, Alexandria B. Boehm

## Abstract

Climate change and urbanization are increasing the distribution of insect vectors of infectious diseases. Dengue virus is an arbovirus that causes nearly 100 million symptomatic infections per year and is endemic in 124 countries, and the range of its mosquito vectors continues to increase. Surveillance of dengue virus infections is complicated by the fact that infections can be asymptomatic and symptoms may not be readily recognizable to clinicians. Here we show that wastewater monitoring can be used to detect dengue virus RNA to yield information about circulation of dengue infections in a community. We collected three samples of wastewater solids per week from three different wastewater treatment plants in Miami-Dade County, Florida where dengue infections have been locally acquired. Using molecular methods, we tested wastewater solids for RNA from the 4 dengue virus serotypes and consistently detected dengue virus 3 RNA at all three wastewater plants, and did not detect the other 3 serotypes. According to publicly available data on dengue infections, the vast majority of infections were caused by serotype 3. Wastewater detection of dengue virus RNA is possible with as few as 4.23 laboratory confirmed dengue cases per 1 million people, based on publicly available infection data.

**Synopsis:** Dengue virus RNA was detected in wastewater solids in a location with local- and travel-associated dengue infections.

## Introduction

Over the last 50 years outbreaks of arthropod-borne viruses (arboviruses) have increased dramatically, due to the limited clinical interventions available, increasing urbanization of tropical countries, and the growing range of their vectors, the *Aedes* and *Culex* mosquitoes.^1–5^ Among important emerging and re-emerging mosquito-borne arboviruses associated with *Culex spp*. (West Nile Virus, Japanese encephalitis) and *Aedes spp*. (Zika virus, yellow fever, chikungunya, dengue virus), dengue virus (DENV) is the most common.^6,7^ In the 1960’s, dengue was endemic in 10 countries and caused a few thousand cases annually, but is now endemic in 124 countries with an estimated 96 million symptomatic infections and 40,000 deaths annually.^4,6^ *Aedes aegypti* is the principal vector for DENV, and infests areas with a total population of more than 3 billion people.^3^ As the distribution of these vectors increases, there is substantial need to monitor the presence and spread of diseases they carry as they enter new communities.

Testing wastewater as a naturally-composited community sample can support infectious disease surveillance, as it contains bodily secretions including urine, feces, vomit, saliva, and sputum. Wastewater testing for viral nucleic acids is rapid and effective for monitoring both the presence and levels of a range of viral diseases, including respiratory viruses (e.g. SARS-CoV-2, influenza), enteric viruses, (e.g. norovirus, rotavirus), and even poxviruses (mpox virus).^8–14^ Wastewater monitoring to capture an emerging viral disease spreading into new geographic areas has been successfully implemented for mpox, and used to supplement public health response to the outbreak.^10^ Early identification of dengue outbreaks is critical for quick and effective action, but traditional approaches including clinical testing, mosquito surveillance, and seroprevalence studies in humans, livestock, and wildlife are resources intensive and often involve reporting delays.^15^ In the United States, existing wastewater monitoring systems in locations with known or anticipated risk of DENV could be leveraged to address this expanding risk.

Wastewater monitoring has been previously proposed as a promising tool for population-level monitoring of DENV, but limited studies testing natural wastewater have not detected the virus.^16,17^ DENV is an enveloped virus in the *Flaviviridae* family with a single stranded RNA genome, and is shed in the urine and saliva of infected individuals and thus likely to be excreted into wastewater.^18–24^ Quantitative data are limited, but shedding in urine has been estimated to be up to around 10^3^ RNA copies/mL (or 10^6^ RNA copies per day) at the peak of shedding, around days 8-10 after symptom onset.^16,19^ Peak shedding occurs during days 0-7 in saliva. DENV shedding in human stool is currently unknown; however, Zika virus, another mosquito-borne virus of the same genus as DENV, has been detected in rectal swabs of infected humans, suggesting DENV might be shed in human stool as well.^25,26^ Laboratory spike-in experiments have demonstrated that DENV can persist in wastewaters for days to weeks.^27–29^

Given the likely feasibility and potential for public health use of wastewater monitoring for dengue, we applied RT-PCR assays for DENV serotypes 1,2,3, and 4 to test wastewater solids from three wastewater treatment plants during summer 2023. These sites, located in a non-endemic area of the United States commonly experiencing both travel-associated and locally-acquired DENV infections, provide a valuable opportunity to assess the feasibility of our approach to identify DENV RNA in wastewater in an area with emerging dengue risk.

## Materials and Methods

### Site and sample collection

Three wastewater treatment plants (WWTPs) in Miami Dade County, Florida were selected: Key Biscayne (KB), North Miami (NM), and South Miami (SM) WWTPs which serve 829,725, 776,150, and 920,528 people, respectively (Figure 1). The county regularly records the highest numbers of dengue cases in the United States outside of non-mainland US territories such as Puerto Rico and the US Virgin Islands.^30,31^ Fifty milliliters of 24-hour composite raw wastewater influent samples were collected using sterile containers by WWTP staff approximately three times per week between 29 June 2023 and 27 September 2023. Samples were sent at 4°C to the laboratory where they were processed immediately.

**Figure 1.**
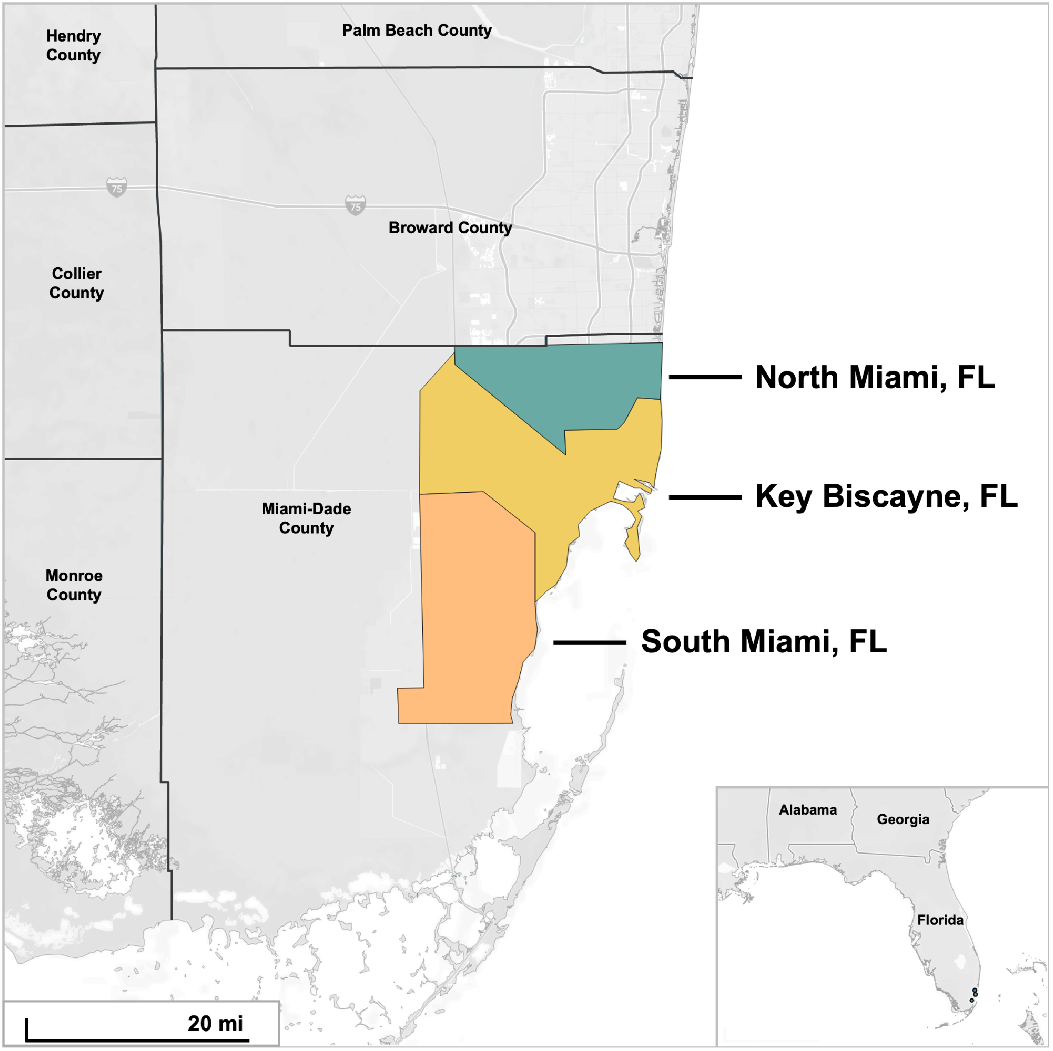
Map of the three sewersheds from which wastewater solids were processed in this study. The border of Miami-Dade County, Florida is shown as well as adjacent counties.

Time between sample collection and receipt at the lab was typically between 1-3 days, during this time, we expect limited degradation of the RNA targets^32^. A total of 112 unique samples were processed. At the lab, the wastewater solids were collected from the influent by settling for 10–15 min, and using a serological pipette to aspirate the settled solids into another tube.

### Assay specificity and sensitivity testing

We used a previous developed set of hydrolysis-probe RT-PCR assays for DENV Types 1, 2, 3, and 4^33^. To ensure their specificity and sensitivity to the intended targets, we tested the assays in silico and in vitro against viral panels and nucleic-acids from the different DENV types (Table 1). For in silico analysis, DENV 1, 2, 3, and 4 genomes were downloaded from NCBI on 6 October 2022.

**Table 1.** Viruses considered in this study and the region of the genome that each assay targeted. Viruses used to test specificity are indicated as “non target testing” and viruses used positive controls are indicated as “target testing”. All non-target controls panels are sold by Zeptomatrix (panels begin with NAT prefix, “Zepto”, Buffalo, NY). ATCC is American Type Culture Collection. ATCC products are synthetic RNA. The NATRVP2.1-BIO panel includes chemically inactivated intact influenza viruses, parainfluenza viruses, adenovirus, rhinovirus, metapneumovirus, and coronaviruses. The NATEVP-C panel includes chemically inactivated intact coxsackieviruses, echovirus, and parechovirus. The full list of species in the panels is available from the vendor.

| Virus | Genomic target | Non-target testing (negatives) | Target testing (positives) |
| --- | --- | --- | --- |
| Dengue Type 1 (DEN-1) | NS5 | NATRVP2.1-BIO<br>NATEVP-C<br>Dengue Serotypes 2, 3, and 4 (ATCC VR-3229SD, VR-3230SD and VR-3231SD) | Dengue Type 1 (ATCC VR-3228SD) |
| Dengue Type 2 (DEN-2) | E gene | NATRVP2.1-BIO<br>NATEVP-C<br>Dengue Serotypes 1, 3, and 4 (ATCC VR-3228SD, VR-3230SD and VR-3231SD) | Dengue Type 2 (ATCC VR-3229SD) |
| Dengue Type 3 (DEN-3) | prM | NATRVP2.1-BIO<br>NATEVP-C<br>Dengue Serotypes 1, 2 and 4 (ATCC VR-3228SD, VR-3229SD, and VR-3231SD) | Dengue Type 3 (ATCC VR-3230SD) |
| Dengue Type 4 (DEN-4) | prM | NATRVP2.1-BIO<br>NATEVP-C<br>Dengue Type 1, 2, and 3 (ATCC VR-3228SD, VR-3229SD, and ATCC VR-3230SD) | Dengue Type 4 (ATCC VR-3231SD) |

### Solids pre-analytical methods

Samples were further dewatered by centrifugation, and dewatered solids were suspended in DNA/RNA Shield (Zymo Research, Irvine, CA) at a concentration of 0.75 mg (wet weight)/ml. The DNA/RNA shield was spiked with bovine coronavirus (BCoV) vaccine as a RNA recovery control. This concentration of solids in buffer has been shown to alleviate inhibition in downstream RT-PCR ^34^. A separate aliquot of dewatered solids was dried in an oven to determine dry weight. RNA was extracted from 6 replicate aliquots of dewatered settled solids suspended in the DNA/RNA Shield, and then it was subsequently processed through an inhibitor removal kit (see SI). The pre-analytical methods are also provided on protocols.io^34^.

### Digital droplet RT-PCR analytical methods

Each replicate RNA extract from each sample (6 per sample) was subsequently processed immediately to measure viral RNA concentrations using digital RT-PCR. We quantified the number of copies of DENV Types 1, 2, 3, and 4 (hereafter, D-1, D-2, D-3, and D-4) using the previously established assays (Table 2). The four assays were run in multiplex using the probe-mixing approach; probes were labeled with one of four fluorescent molecules as follows: D-1 (FAM, 6-fluorescein amidite), D-2 (FAM), D-3 (ROX, carboxyrhodamine), and D-4 (ATTO590) except that between 27 July and 27 September 2023, D-1 was labeled with HEX. Note this means that concentrations of D-1 and D-2 are only available in aggregate until 27 July 2023; as discussed below, this is inconsequential since all values were non-detect for these targets. We also measured concentrations of pepper mild mottle virus (PMMoV) RNA; PMMoV is highly abundant in wastewater globally^35^ and is used as an internal recovery and fecal strength control^36^. We also measured concentrations of BCoV RNA. Further details of the RT-PCR including cycling conditions, ingredients, replication, positive and negative controls, as well as thresholding, are in the SI. Primer/probe sequences are in Table 2. In order for a sample to be recorded as positive, it had to have at least 3 positive droplets.

**Table 2.**
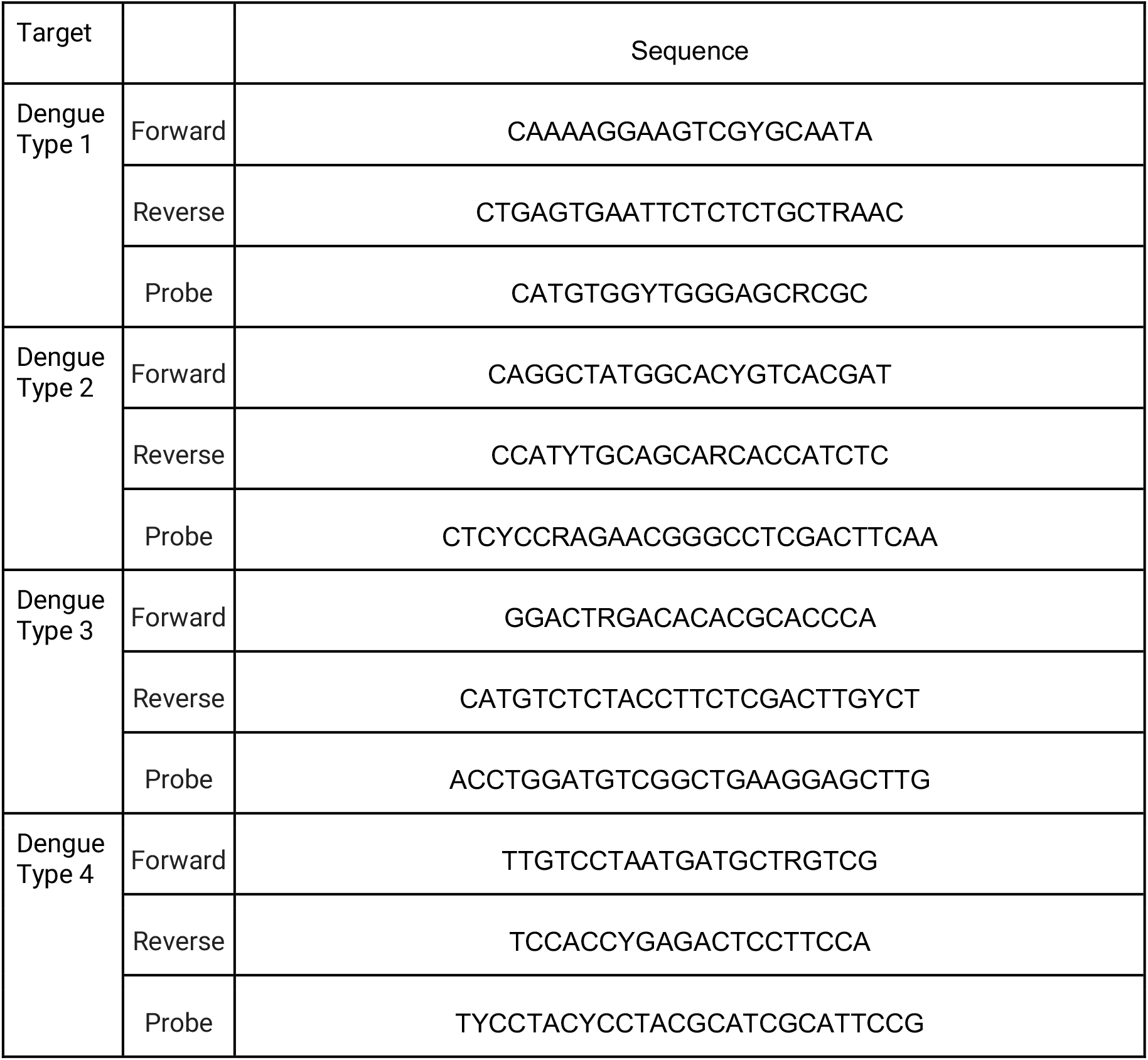
Primer and hydrolysis probes targeting different Dengue serotypes. The primers and probes were published by Santiago et al.^33^ as reported in Table 1 labeled as “CDCDENV-1– 4RealTimeRT-PCR”. Each probe contained a fluorescent molecular (FAM, HEX< ROX, or ATTO590), as well as ZEN, a proprietary internal quencher from IDT; and IBFQ, Iowa Black FQ. Note that unit 26 July 2023, Dengue Type 1 probe was used with a FAM molecule instead of HEX.

Concentrations of RNA targets were converted to concentrations per dry weight of solids (copies per gram dry weight (cp/g)) using dimensional analysis. The error is reported as standard deviations and includes the errors associated with the Poisson distribution and the variability among the replicates. Three positive droplets across 10 merged wells corresponds to a concentration ∼500 cp/g for solids, respectively). Concentrations of DENV RNA in the samples are available through the Stanford Digital Repository (https://doi.org/10.25740/bn739zz5683).

### Dengue Case Surveillance

Data for dengue case detection and subtyping was obtained from the Florida Health Department weekly arbovirus report, which are publicly available.^37^ Dengue is a reportable disease in Florida, and practitioners are required to report upon suspicion and ordering of laboratory tests.^38^ Both confirmed and probable dengue cases are counted in Florida residents and non-resident visitors to the state. Serotyping is based on PCR-confirmation.

State- and county-aggregated incident case data were digitized from the weekly reports for the study period (18 June 2023 - 30 September 2023; MMWR weeks 25-39). Data on the specific DENV serotype responsible for each incident case are available for only state-aggregated, travel-associated cases, and county-aggregated locally-acquired cases.

## Results and Discussion

### QA/QC

Previously designed DENV assays for four DENV serotypes were found to be both specific and sensitive, able to detect their intended targets with no cross reactivity. Negative and positive extraction and PCR controls on all plates used for environmental sample testing were negative and positive. BCoV recoveries and PMMoV values indicate median recoveries close to 100%, and stable PMMoV between and within WWTPs (see supporting information, SI). This suggests DENV RNA concentrations can be compared over time and between WWTPs as they have similar high recoveries and consistent fecal strength. Additional details related to the Environmental Microbiology Minimal Information (EMMI) guidelines are in the SI (Table S1).

When these assays were applied prospectively to samples of wastewater solids obtained from three WWTPs in a location in the United States with increasing concerns about local dengue transmission, we found that DENV serotype 3 (DENV-3) RNA was consistently detected during a time period when both travel-associated and locally-acquired cases of dengue were identified in the county (Figure 2). DENV-3 RNA was detected in 24 of 112 (21%) samples from all three WWTP sites. DENV-3 RNA concentrations ranged from below the detection limit (∼500 copies(cp)/gram wastewater solids) to a maximum of 4.1x10^3^ cp/g (median = nondetect). All wastewater samples were non-detect for DENV-1, -2, and -4 RNA. During the study period, there was at least one sample positive for DENV-3 from a Miami-Dade site from 13 of 14 weeks (Figure S1,). A description of sampling dates and rates of detection for individual sites are available in Table S2.

**Figure 2.**
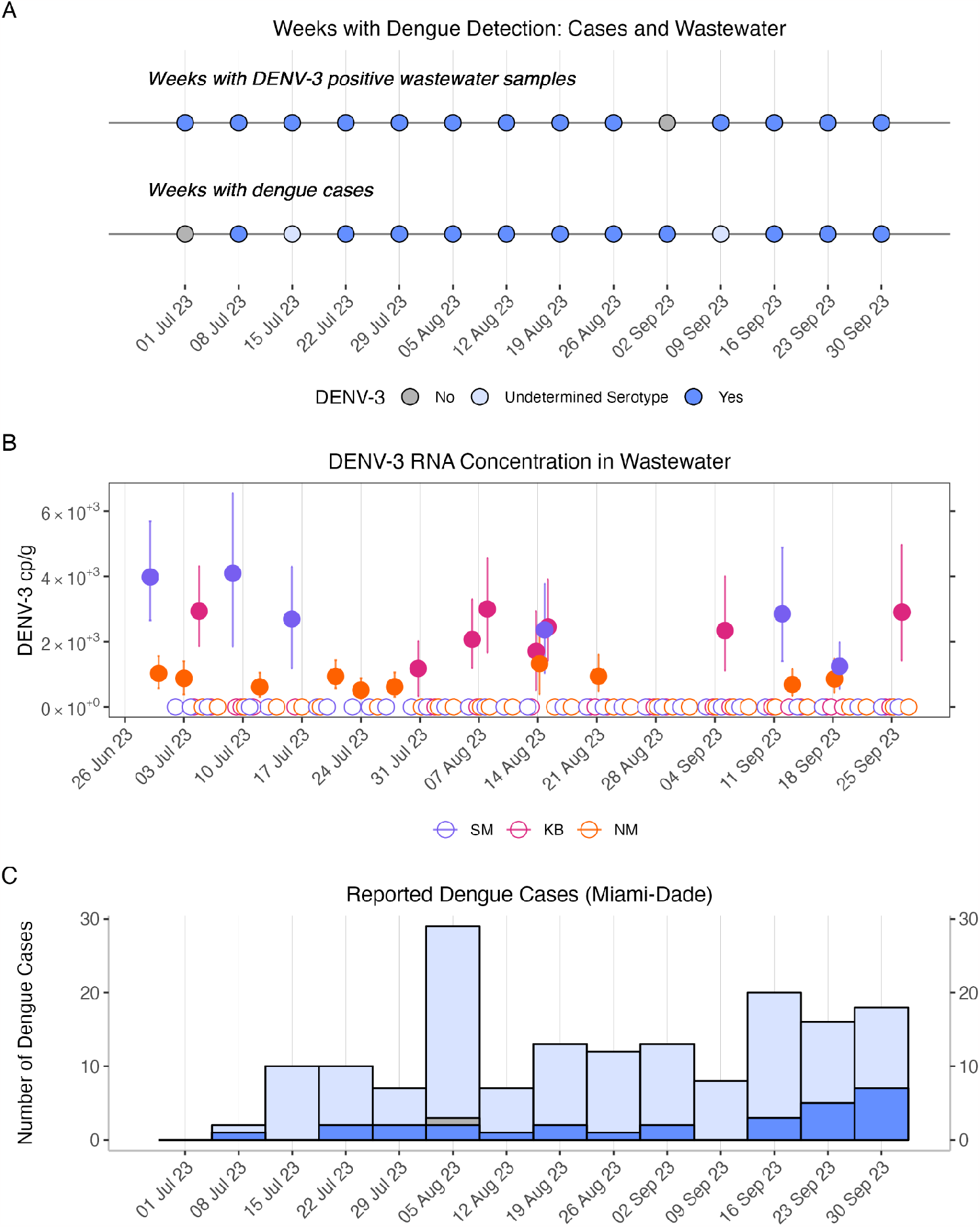
Detection of DENV-3 RNA in wastewater and reported dengue cases in Miami-Dade County. A: Weeks during the study period with detection of DENV-3 in wastewater at any of the three sites (top) and weeks with dengue cases identified in Miami-Dade county (bottom) B: Concentrations of DENV-3 RNA in each sample processed in this study. Errors represent standard deviations on the measurement. KB is Key Biscayne, NM is North Miami, and SM is South Miami WWTPs. C: Dengue cases reported in Miami-Dade county, from Florida Arbovirus Weekly Reports. Overall bar height represents the total number of cases per MMWR week for both travel-associated and locally-acquired cases. Colors represent available information on serotyping. Locally-acquired cases were serotyped and confirmed DENV-3 cases are shown in dark blue and other serotypes in grey; the remaining travel-associated cases where serotype was not provided are shown in light blue.

Although the three WWTPs do not cover the entire geographic area of the county, they are estimated to serve 95% of the county’s population and therefore it is likely that most recorded cases are within the sampled sewersheds (2.5 million people served, total Miami-Dade population 2.6 million). During the study period, there were 165 incident cases of DEN reported in Miami-Dade County (132 travel-associated, 33 locally acquired). Serotyping information was not provided for travel-associated cases in the county, however 71% (166/234) of travel-associated cases state-wide were serotyped as DENV-3. For the 33 locally-acquired cases of dengue, 28 (85%) were positively identified as DENV-3. One case was positively identified as DENV-2 (3%) and the other four cases were of unknown serotype (12%). The median number of incident dengue cases per week was 11 (range = 0-29), and median incident cases with a confirmed DENV-3 serotype was 2 (range = 0-7). Although serotype was not provided for the majority of cases, rates of DENV-3 statewide suggest it accounts for the majority of cases.

Depending on whether only confirmed DENV-3 or all dengue cases are considered, wastewater results show detection of DENV-3 in a population with an estimated weekly incidence rate of 0.77 - 4.23 cases/ 1 million people, respectively.

Because a large proportion of people infected with DENV for the first time may be asymptomatic or mildly symptomatic and not seek care, and because the disease is rare in the United States and may not be easily recognizable to clinicians, dengue surveillance is not expected to capture all cases.^6,39,40^ Therefore the case surveillance data described here is likely an underestimate of both travel-associated and locally-acquired infections, and may also be incomplete due to reporting delays.^41^ Nonetheless, results show that DENV-3 RNA is detectable in wastewater in an emerging region with a low incidence rate and limited local transmission.

Wastewater has been considered to supplement DENV surveillance in part due to a focus on sentinel surveillance to identify nascent outbreaks, as the success of mitigation efforts, including vector control, depends on rapid response.^15,42,43^ Current dengue surveillance includes both case reporting, which will not capture individuals who are not diagnosed due to subclinical infections or missed diagnosis, and mosquito surveillance, which is time and resource intensive. A recent meta-analysis suggests that the prevalence of asymptomatic dengue infections overall was 59.26% (95% CI: 43.76-74.75, I2 = 99.93%), and other work shows that they are infectious.^39,40^ Despite documented cases of local transmission during the study time period, concurrent mosquito surveillance in Miami-Dade county showed consistent capture of *Ae. aegypti* without any mosquito testing positive for DENV [personal communication, Dr. Isik Unlu, Miami-Dade County]. In cases like this, wastewater provides an opportunity to sample at the population level to supplement and sustain sentinel surveillance for DENV in human populations by indicating the likely presence of human cases.^8,9^ Note that while there is some chance that mosquitoes could appear in wastewater and contribute to detection, this would still be a valid indicator of community presence of the disease (although DENV is not transmissible to humans through wastewater, as it requires the mosquito vector).

This work suggests that monitoring for DENV in wastewater is feasible at low levels of incidence and in emerging regions, and therefore could be a valuable addition to dengue surveillance programs. Previous studies have not successfully detected DENV in natural wastewaters^17^. A difference in this study is the use of solids, many previous studies focus on analysis of the liquid phase. Other work has shown that many viruses are found in concentrations several orders of magnitude higher in wastewater solids^44–48^. To better understand and utilize results, more work is needed to characterize DENV shedding and persistence and partitioning in wastewater and to characterize the relationship between measured concentrations of DENV RNA in wastewater and infections in the community. Current wastewater methods cannot distinguish between locally-acquired and travel-associated cases, information that is critical to understanding the scope of outbreaks and risk to the community.

## Data Availability

Data are available at the Stanford Digital Repository.

https://doi.org/10.25740/bn739zz5683

## Supporting Information

Additional methodological details as well as Tables S1-S2 and Figures S1-S3.

## Acknowledgements

We acknowledge the staff at the three Miami-Dade County wastewater treatment plants for sample collection, as well as local health department professionals Isik Unlu, Samir Elmir, Yesenia Villalta, Ian Stryker, Andrea Morrison, and Daneille Stanek for their feedback on and contributions to the project.

