## Supporting Information for "Wastewater detection of emerging arbovirus infections: Case study of Dengue in the United States"

**Number of pages: 9**

**Number of Tables: 2**

**Number of Figures: 3**

**RNA extraction and purification.** RNA extraction and purification was done using the Chemagic Viral DNA/RNA 300 kit H96 for the Perkin Elmer Chemagic 360 (Perkin Elmer, Waltham, MA). It was followed by PCR inhibitor removal with the Zymo OneStep-96 PCR Inhibitor Removal kit (Zymo Research, Irvine, CA). 300 µl of the suspension entered into the nucleic-acid extraction process and 50 µl of nucleic-acids are retrieved after the inhibitor removal kit.

**Further details of RT-PCR.** Ten replicate wells were run for D-1, D-2, D-3, and D-4 each sample, using the 6 replicate nucleic-acid extracts - 4 of the 6 were chosen to run in duplicate. Two replicate wells were run for each sample for PMMoV and BCoV in duplex, as described by Boehm et al.[34] Extraction negative (BCoV spiked buffer, 3 wells) and positive (buffer spiked with positive control cDNA of targets, 1 well) controls, and PCR negative (molecular grade water, 3 wells) and positive controls (cDNA, 1 well) were run on each 96 well plate.

ddRT-PCR was performed on 20 µl samples from a 22 µl reaction volume, prepared using 5.5 µl template, mixed with 5.5 µl of One-Step RT-ddPCR Advanced Kit for Probes (Bio-Rad 1863021), 2.2 µl of 200 U/µl Reverse Transcriptase, 1.1 µl of 300 mM dithiothreitol (DTT) and primers and probes mixtures at a final concentration of 900 nM and 250 nM respectively. Primer and probes for assays were purchased from Integrated DNA Technologies (IDT, San Diego, CA) (Table 2, main text). DENV RNA was measured in reactions with undiluted template whereas PMMoV and BCoV assays were run in duplex on template diluted 1:100 in molecular grade water.

Droplets were generated using the AutoDG Automated Droplet Generator (Bio-Rad, Hercules, CA). PCR was performed using Mastercycler Pro (Eppendforf, Enfield, CT) with the following cycling conditions: reverse transcription at 50 °C for 60 min, enzyme activation at 95 °C for 5 min, 40 cycles of denaturation at 95 °C for 30 s and annealing and extension at 59 °C (for DENV assays) or 56 °C (for PMMoV/BCoV) for 30 s, enzyme deactivation at 98 °C for 10 min then an indefinite hold at 4 °C. The ramp rate for temperature changes were set to 2 °C/second and the final hold at 4 °C was performed for a minimum of 30 min to allow the droplets to stabilize. Droplets were analyzed using the QX200 or the QX600 Droplet Reader (Bio-Rad). A well had to have over 10,000 droplets for inclusion in the analysis. All liquid transfers were performed using the Agilent Bravo (Agilent Technologies, Santa Clara, CA).

Thresholding was done using QuantaSoft Analysis Pro Software (Bio-Rad, version 1.0.596) and QX Manager Software (Bio-Rad, version 2.0). In order for a sample to be recorded as positive, it had to have at least 3 positive droplets. Replicate wells were merged for analysis of each sample.

**BCoV Recovery and PMMoV values.** Median (interquartile range, IQR) BCoV recoveries across all samples were 1.0 (0.8, 1.3) for KB, 0.9 (0.7, 1.1) for NM, and 0.9 (0.8, 1.1) for SM. Median (IQR) PMMoV were $1.5 \times 10^8$ ($1.2 \times 10^8$ - $1.8 \times 10^8$) cp/g for KB, $1.1 \times 10^8$ ($0.8 \times 10^8$ - $1.8 \times 10^8$) cp/g for NM, and $1.6 \times 10^8$ ($1.3 \times 10^8$ - $2.3 \times 10^8$) cp/g for SM (Figure S2).

**Additional details related to the EMMI guidelines.** Eighteen (18) wastewater samples from the study were selected at random for this analysis; this represents 15% of the samples processed in the study. The average (standard deviation) number of partitions (droplets) for the across the 10 replicate wells was 177,871 (21,959) for the reaction for DEN assays. The volume of the partitions, as reported by the machine vendor is 0.00085 µL. The mean and standard deviation of copies per partition for each DEN target is shown in Table S1. An example fluorescent plot from the QX600 (6 color reader), as well as a spreadsheet version of the EMMI checklist (Figure S3) is included in the Stanford Digital Repository with the deposited data (https://doi.org/10.25740/bn739zz5683).

Table S1. Additional details related to the EMMI[1] guidelines. For each target measured in this study, the mean and standard deviation (sd) of the total number of copies of target per partition. Abbreviations for the targets are provided in the main text.

| Target | D1 | D2 | D3 | D4 |
| --- | --- | --- | --- | --- |
| mean | $6.8x10^{-7}$ | $3.2x10^{-7}$ | $5.7x10^{-6}$ | $1.7x10^{-6}$ |
| sd | $1.9x10^{-6}$ | $1.3x10^{-6}$ | $5.2x10^{-6}$ | $2.9x10^{-6}$ |

Table S2. Information about samples collected at the three wastewater treatment plants in Miami-Dade Florida. Date range is the date range during which samples were collected. N is total samples collected, # pos is number positive for DENV-3, Min, Median, and Max Concentration are the minimum, median and maximum concentration in units of copies per gram dry weight solids.

| WWTP | Date Range | N | # pos for DENV-3 | Min, Median, and Max Concentration |
|---|---|---|---|---|
| NM | 6/30/2023 - 9/27/2023 | 39 | 10 | Min = nondetect<br>Med = nondetect<br>Max = $1.32 \times 10^3$ |
| KB | 7/4/2023 - 9/26/2023 | 34 | 8 | Min = nondetect<br>Med = nondetect<br>Max = $3.00 \times 10^3$ |
| SM | 6/29/2023 - 9/26/2023 | 39 | 6 | Min = nondetect<br>Med = nondetect<br>Max = $4.09 \times 10^3$ |

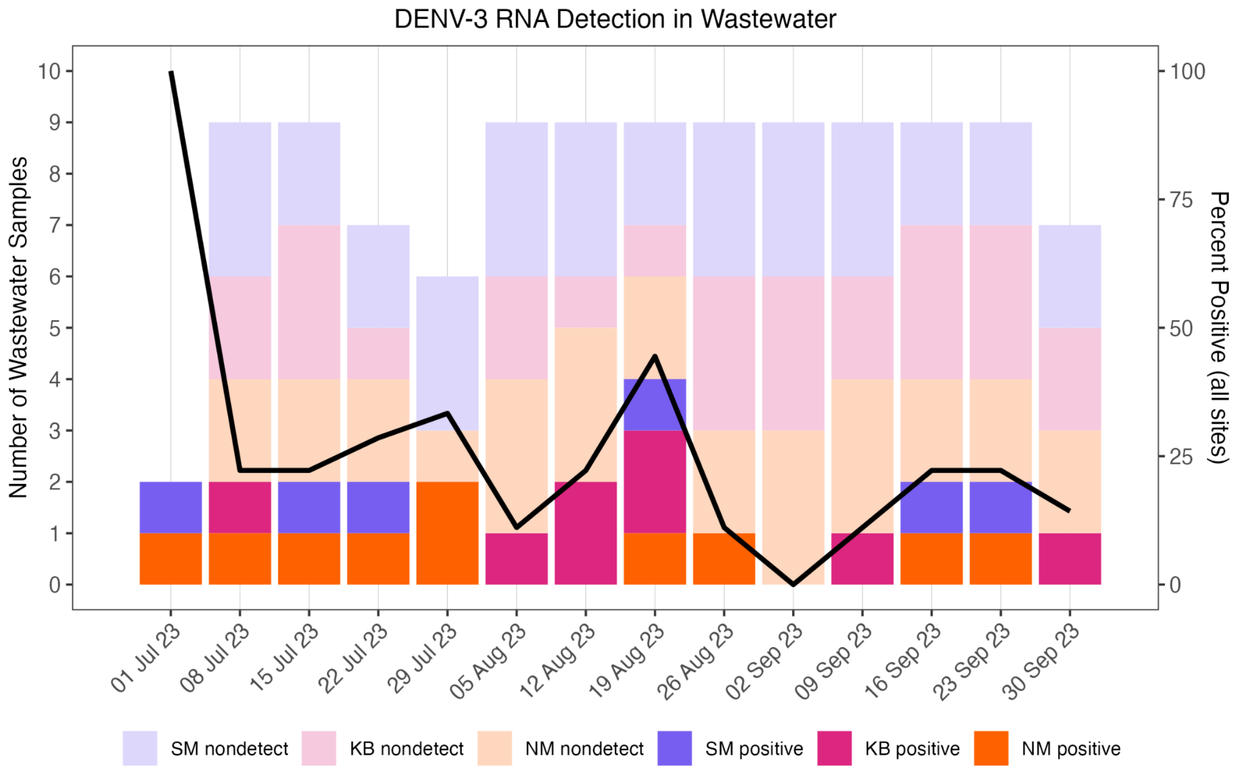

Figure S1.  Wastewater results by week for DENV-3 by site. Left axis: bars represent total samples positive for DENV-3 RNA and detected by site; colors indicate the site associated with each samples. KB is Key Biscayne, NM is North Miami, and SM is South Miami WWTPs as shown in the main text Figure 1. Right axis: The black line represents the percentage of samples testing positive in each week, across all sites.

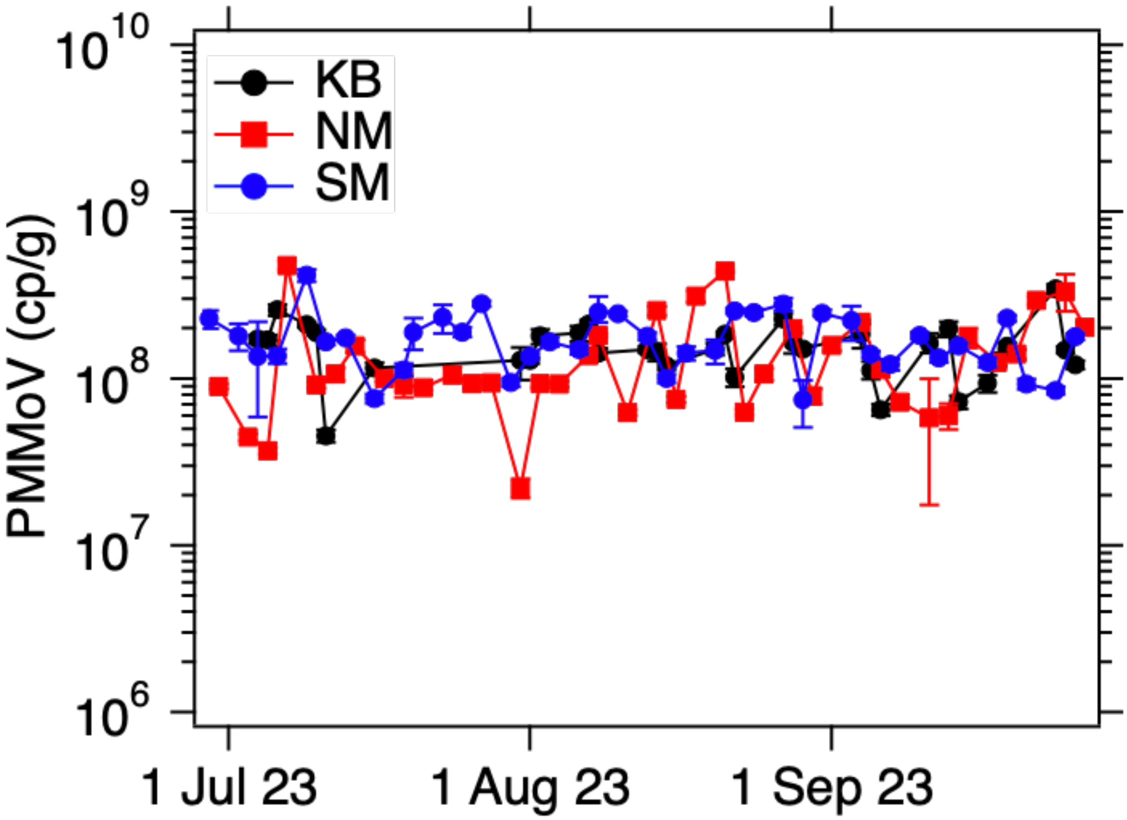


Figure S2. Concentrations of PMMoV in each sample processed in this study. Errors represent standard deviations on the measurement. KB is Key Biscayne, NM is North Miami, and SM is South Miami WWTPs as shown in the main text Figure 1.

### Study Description

| Study Description | Environmental Sampling | Sample Treatment | Sample Reduction | Nucleic-acid Extraction | Reverse Transcription | PCR Amplification | Analysis |
|---|---|---|---|---|---|---|---|
| Dengue Virus in Wastew<br>Sep-23<br>Alexandria Boehm | Notes: Described in methods section. Same samples used in other publications which are sited | Notes: None | Notes: Resuspension of solids in a buffer, described in methods section | Notes: Described in methods section | Notes: Described in methods | Notes: Provided in the methods section. | Notes: Details of analysis provided in the methods section. |

### Control Checklist

| Control Checklist | Environmental Sampling | Sample Treatment | Sample Reduction | Nucleic-acid Extraction | Reverse Transcription | PCR Amplification | |
|---|---|---|---|---|---|---|---|
| Step performed | ☑ | ☐ | ☑ | ☑ | ☑ | ☑ | |
| Step has control info | ☐ | ☐ | ☐ | ☑ | ☑ | ☐ | Negative controls |
| # of control replicates | 0 | | 0 | 3 | 3 | na | |
| Control result reported | ☐ | ☐ | ☐ | ☑ | ☑ | ☐ | |
| Method for handling failed controls described | ☐ | ☐ | ☐ | ☑ | ☑ | ☐ | |
| Step has control info | ☑ | ☐ | ☐ | ☑ | ☑ | ☐ | Positive controls |
| Control identity described | ☑ | ☐ | ☐ | ☑ | ☑ | ☐ | |
| Control quantification method described | ☑ | ☐ | ☐ | ☑ | ☑ | ☐ | |
| # control replicates | endogenous | | 0 | 1 | 1 | na | |
| Control result reported | ☑ | ☐ | ☐ | ☑ | ☑ | ☐ | |
| Method for handling failed controls | ☑ | ☐ | ☐ | ☑ | ☑ | ☐ | |

### Process checklist

| Environmental Sampling | |
|---|---|
| Sample procedure | ☑ |
| Number of samples | ☑ |
| Sample amount, mean, range | ☑ |
| Sampling locations, dates, times | ☑ |
| Sample storage conditions | ☑ |
| **Sample Treatment** | |
| Treatment procedure | ☐ |
| Reagents | ☐ |
| **Sample Reduction** | |
| Reduction procedure | ☑ |
| Reagents | ☐ |
| Concentration factor | ☑ |

| Nucleic-acid Extraction | |
|---|---|
| Extraction procedure | ☑ |
| Volume or mass extracted, volume or mass obtained | ☑ |
| Extract storage conditions | ☑ |
| **Reverse Transcription** | |
| One- or two-step | ☑ |
| cDNA storage conditions (if 2 step) | ☐ |
| Reaction temperatures and times | ☑ |
| Reaction reagents and concentrations | ☑ |
| Priming method | ☑ |
| Reaction volume, added template amount | ☑ |
| RT efficiency assessment procedure (if 2-step) | ☐ |
| RT control description (if two-step) | ☐ |
| RT efficiency reported (if 2-step) | ☐ |

| qPCR or dPCR | |
|---|---|
| Target gene name, amplicon length | ☑ |
| Thermocycling temp and times | ☑ |
| Master mix composition: vendors, concentrations | ☑ |
| Additives: vendors, composition | ☑ |
| Template amount added, pre-treatment (if any) | ☑ |
| Primers: sequences, concentrations, vendors, references | ☑ |
| Amplicon confirmation method (probe, melt curve details, etc) | ☑ |
| Probe sequence, concentration, vendor, reference | ☑ |
| Instrumentation | ☑ |
| Inhibition assessment procedure | ☑ |
| Inhibition control description (if used) | ☐ |
| Number of samples tested and found inhibited | ☐ |

| Analysis - dPCR | |
|---|---|
| Threshold settings | ☑ |
| Technical replicates, number, well merging | ☑ |
| Partitions measured, number, mean, variance | ☑ |
| Partition volume | ☑ |
| Target copies per partition, mean, variance | ☑ |
| Program used for dPCR analysis | ☑ |
| Explanation of control results, example plots | ☑ |
| **Analysis - qPCR** | |
| Technical replicates, number, calculations | ☐ |
| Calibration standards, description, source | ☐ |
| Method of quantifying standards | ☐ |
| Calibration curve slope | ☐ |
| Calibration curve R2 | ☐ |
| Lowest standard measured or 95% LOD | ☐ |
| Cq value determination methods | ☐ |

Note to users: This checklist is provided as guidance for best practices for reporting, but is not meant to be prescriptive. Not all items in the check list will apply to all studies. Please see Borchardt et al. The

Version 2.0

Figure S3. The EMMI [1] checklist. A spreadsheet version of the checklist is available at the Stanford Digital Repository (https://doi.org/10.25740/bn739zz5683).
